# Predisposing, enabling, and need factors influencing rapid uptake of the World Health Organization-endorsed TB diagnostic technologies in Africa

**DOI:** 10.1101/2024.03.01.24303364

**Authors:** Jean de Dieu Iragena, Achilles Katamba, Anandi Martin, Moses Joloba, Willy Ssengooba

## Abstract

**Background:** The implementation of rapid tuberculosis (TB) diagnostics is essential for TB control. Factors influencing their uptake in Africa are unknown. We conducted a survey to collect the status and document Predisposing, Enabling, and Need (PEN) factors influencing so that we understand the associated barriers and inform interventions to improve the uptake.

**Methods:** We designed, piloted, and sent out a survey questionnaire in January 2023 to the National TB Programme (NTP), and National TB Reference Laboratory (NTRL) managers and key partners of the Ministry of Health in the 47 Member States of the World Health Organization African Region (WHO/AFR). Responses were accepted until July 2023. We performed qualitative and quantitative data analysis using STATA version 14.0.

**Results:** From the 47 eligible countries, 22 responses (47%) were received from the NTRL managers, 17 (36%) from Technical Assistants (TAs) for NTRL and NTP, and 8 (17%) from the NTP managers. Our findings showed that it took between two to nine years from the endorsement of a new technology and its full implementation and the years increased with increasing test complexity. Competence of staff and laboratory preparedness were the main predisposing factors; availability of funds was the main enabling factor whereas the increase in TB incidence and mortality as well as the emergency of MDR-TB were the key need factors. Good Governance and political commitment aligned with the existence of the Directorate of Laboratory Services and the NTRL were key facilitators to drive the adoption, adaptation, and implementation.

**Conclusion:** Our findings demonstrated that the uptake of TB diagnostics in Africa is slow. Taking into account the competence of staff, the availability of funds, and the burden of TB as the main PEN factors identified respectively could help in speeding up the uptake and rapid implementation of any new technology.

**Key questions:** *What is already known:* The rapid implementation of TB diagnostic technologies is important for the TB control programme. Every year, millions of TB presumptive individuals go undiagnosed and therefore miss treatment and continue to spread the infection.

*What are the new findings?:* Despite the availability of TB diagnostic technologies and WHO guidelines for use, our study reveals that it takes several years before countries can fully implement and report the impact of the use of a diagnostic technology after its endorsement. Staff competence, the availability of funds, and the TB incidence, and mortality as well as the emergency of MDR-TB are strong predisposing, enabling, and need factors influencing the uptake of a newly WHO-endorsed TB diagnostic respectively. To embrace these factors, laboratory governance is a core facilitating factor in implementation.

*What do the new findings imply?:* Delays in the uptake of a new diagnostic technology may perhaps explain the rationale behind the million people missing TB diagnosis and thereby treatment every year. Country-led Laboratory Governance is an important solution to improve the implementation and allow a quick access to diagnostic while addressing PEN factors.

## BACKGROUND

The WHO’s End Tuberculosis (TB) Strategy^1^ calls for the early diagnosis of TB including universal drug-susceptibility testing (DST) and a quality-assured laboratory network.^2^

In 2022, an estimated 10.6 and 2.5 million people fell ill with TB globally and in Africa respectively, with almost 1.7 million (68%) of the new and relapse TB cases being diagnosed and notified leaving a gap of 32% undiagnosed and unnotified.^3^ WHO has endorsed different diagnostic technologies since 2007 to date with advantages and limitations.^4^ In 2007, a highly sensitive MGIT liquid culture system was endorsed as a first-line culture method instead of Lowenstein–Jensen (LJ) solid medium culture.^5^ Then, WHO endorsed the molecular GenoType®MTBDR*plus* and GenoType®MTBDR*sl* (Hain Lifescience, Nehren, Germany) for rapid detection of RIF, INH resistance,^6^ and second-line fluoroquinolone (SL-FQ) and injectable agents (IAs) respectively.^7^ Nevertheless, their use has not yet solved the diagnostic dilemma of low uptake, largely due to the need for expensive laboratory infrastructure with proper design, extensive biosafety precautions to avoid cross-contamination, and specialized staff ^8^.

In 2010, WHO endorsed the use of Light-emitting diodes (LED) microscopy ^9^ being 10% more sensitive than conventional light microscopy (LM) using Ziehl-Neelsen (ZN) staining with a positive impact in the HIV population^10^. Then another Policy on rapid Non-commercial culture and DST methods for screening patients at risk for MDR-TB, was endorsed ^11^ but because of technical complexity, and sophisticated laboratory infrastructure, the use of these techniques has been limited ^12^.

Late in 2010, The Xpert®MTB/RIF assay (Xpert) (Cepheid, Sunnyvale, CA, USA) was endorsed as a breakthrough, rapid, and more sensitive technology but limited to MDR or HIV associated TB ^13^ and in 2013, a new policy on its use on all people suspected of TB was made as an advantage.^14^ The test was upgraded in 2017 to a better version with improved performance in the detection of TB and RIF resistance, the Xpert®MTB/RIF Ultra assay (Ultra) ^15^ ^16^. Recently in 2019, a new class of Xpert®MTB/XDR cartridge to detect susceptibility to INH, ethionamide, FQs, and IAs (amikacin, kanamycin, and capreomycin) has been recommended^17^. In contrast to the Xpert MTB/RIF and Ultra, the test can only run on a 10-color machine, hence the swap from 6-color by the manufacturer should be encouraged.^18^ The running cost, the infrastructure, equipment maintenance, and skilled personnel are the most noted limitations for the optimal utilization of Xpert in general.

In 2015, a policy on Lateral Flow Lipoarabinomannan (LF-LAM), a rapid diagnostic test (Alere Determine^TM^TB LAM Ag, Alere Inc, Waltham, MA, USA) as point-of-care test for the diagnosis and screening of active TB in urine of people living with HIV (PLHIV) was issued ^19^ and in 2019, its use in the diagnosis of active tuberculosis in PLHIV was made.^20^ Despite this and the test does not require laboratory infrastructure and skilled personnel, the adoption and uptake of the test have been slow.^21^ New evidence has emerged with the Fujifilm SILVAMP TB LAM, Tokyo, Japan (FujiLAM) – more sensitive and eligible in a larger proportion of hospitalized and non-hospitalized patients.^22^ However, lot-to-lot variability that affected sensitivity and specificity estimates limited utilization.^23^

In 2016, TB LAMP [the Loopamp™ *Mycobacterium tuberculosis* complex (MTBC) detection kit, Eiken Chemical Company] for use as a rapid alternative to sputum-smear microscopy was endorsed ^24^. The test is rapid, robust and requires the same biosafety level as microscopy, however, it does not provide a resistance profile, hence its use was limited. In 2020, a new molecular TB diagnostic tool named Truenat (MTB, MTB Plus, and MTB-RIF) was developed by Molbio Diagnostics, Bangalore, India for the detection of TB and RIF-resistance from sputum samples within an hour, and Truenat MTB identified more positives among culture-confirmed samples than Xpert and had higher sensitivity.^25^

Despite all these technologies being in place for several years, their reported uptake in high TB burden countries remains low and the factors influencing their uptake are unknown. Our study aimed at documenting the status of the uptake and PEN factors influencing the uptake of WHO-endorsed TB diagnostic technologies in the 47 member states of WHO/AFR considering TB diagnostic technologies endorsed by WHO between 2007-2021. The identified factors will help to inform rapid TB diagnostic uptake and roll-out of the already endorsed ones and in the pipeline.

## METHODS

There was a data collection survey questionnaire designed in French and English languages to collect information on the status of the uptake and document PEN factors influencing the uptake of the WHO-endorsed TB diagnostic technologies. The questionnaire was submitted together with the study protocol to the Research and Ethics Committee of the School of Biomedical Sciences (SBS-2022-154) at the College of Health Sciences, Makerere University, Kampala, Uganda, and to the Uganda National Council for Science and Technology (HS2393ES) for review and approval before it is converted to an online survey.

### Data collection

An online survey questionnaire was designed, constructed from the REC-approved version, and piloted using the Google tool (https://docs.google.com/forms/u/0/) (**Supplementary file 1**). This was sent in January 2023 to NTP Managers, NTRL Managers, key partners, and officials from the Ministry of Health (MoH) in all 47 countries of the WHO/AFR (**Figure S1**). The cover page of the survey questionnaire had an informed consent section with participant information and agreement for willingness to participate in the survey. A follow-up by phone and/or email as a reminder in case of a delay in responding was made.

### Survey questionnaire

This consisted of a list of questions made in a way participants could respond to multiple-choice questions and/or opened answer questions, among others. Briefly: the questionnaire focused on the country’s situation at different periods regarding national security, policy reform, the existence of national laboratory policy including the existence of the Directorate of Laboratory Services (DLS) within the MoH, whether countries have formally assigned NTRL as stand-alone or integrated into the National Public Health Laboratory (NPHL), among others. Participants were required to report whether the TB Laboratory Strategic Plan exists, as a stand-alone, or integrated into the National TB Strategic Plans to better understand how important it is budgeted. The information on the formal collaboration between the NTRL and the WHO-TB Supranational Reference Laboratory (SRL) was collected. The status of funding mechanisms and technical assistance to countries was a key driver that may influence the rapid or slow update. Factors influencing the uptake of each TB diagnostic technology after endorsement by WHO were collected starting in 2007.

Participants were also required to respond, per each technology, to the implementation status according to the 3 defined phases: laboratory preparedness, technology transfer, and routine testing. Each phase was given 3 years with the expectation to get more countries starting the routine testing at least 6 years after the recommendation was issued. The Molecular WHO-recommended Rapid Diagnostics (WRDs) was given less than 6 years as the laboratory preparedness and technology transfer phases need minimum requirements. The exception was for LPA for which the laboratory preparedness phase required laboratory infrastructure, biosafety, and trained human resources to be addressed before the technology could be transferred to countries. Responses were analyzed by looking at the laboratory Governance including DLS, Laboratory policy, laboratory strategic plans with assigned budget, formal NTRL to lead the TB laboratory capacity building, and each endorsed test including its uptake across all three 3 phases of the implementation.

Data concerning the status of diagnostic uptake and PEN factors including barriers related to them were collected. PEN factors were not collected for LED microscopy replaced by molecular WRDs as initial TB diagnostics, rapid non-commercial culture, and DST methods as their uptakes were very low and Xpert MTB/RIF being replaced by Ultra

### Data management and analysis

Responses were summarized in variables for analysis. Multiple responses were managed to remain with one response per country following the criteria as follows: 1) Responses from NTRL Managers were considered a first priority and more representative since the study subject is about laboratory diagnostics. 2) Where NTP and NTRL Managers both gave a complete response, we reviewed their responses, and where there were discrepancies, we got back to them to clarify and consider their final opinion to select the most knowledgeable among both. 3) We considered the survey’s completeness, and the responder’s experience in position, and merged responses between respondents per country. 4) Where responses were discordant and/or missing, verifying the right response to retain was considered. Responses were accepted until July 2023. We performed a qualitative analysis in a logistic model using STATA version 14.0

We analyzed responses by mapping countries as rapid, moderate/slow implementers in color-coded categories (Green and Orange colors). Rapid implementers were assigned a green color and moderate/slow implementers an orange color if the specified technology shows its implementation occurred within 1 to 3 years and 3 to 5 years onward respectively.

We analyzed the status of uptake per technology and PEN factors influencing the implementation. This took into account the enabling environment such as 1) Laboratory Governance and structure within the MoH including influencers of the uptake and funders, 2) Policy adoption and their implementations with different phases of laboratory preparedness, technology transfer, and the onset of routine testing and 3) PEN factors influencing the uptake.

## RESULTS

A total of 68 responses were received from all the countries of which 40 were from both NTP and NTRL Managers, 28 (41.2%) were from the NTRL Managers only, 28 (41.2%) from Technical Assistants (TAs) for NTRL and NTP, and 12 (17.7%) from NTP Managers only. After removing duplicates, less experienced respondents, and incomplete responses, 47 responses were considered, out of which 22 (47%) were from the NTRL Managers, 17 (36%) were from TAs to NTRL and NTP, and 8 (17%) were from the NTP Managers (**Figure 1**).

**Figure 1.**
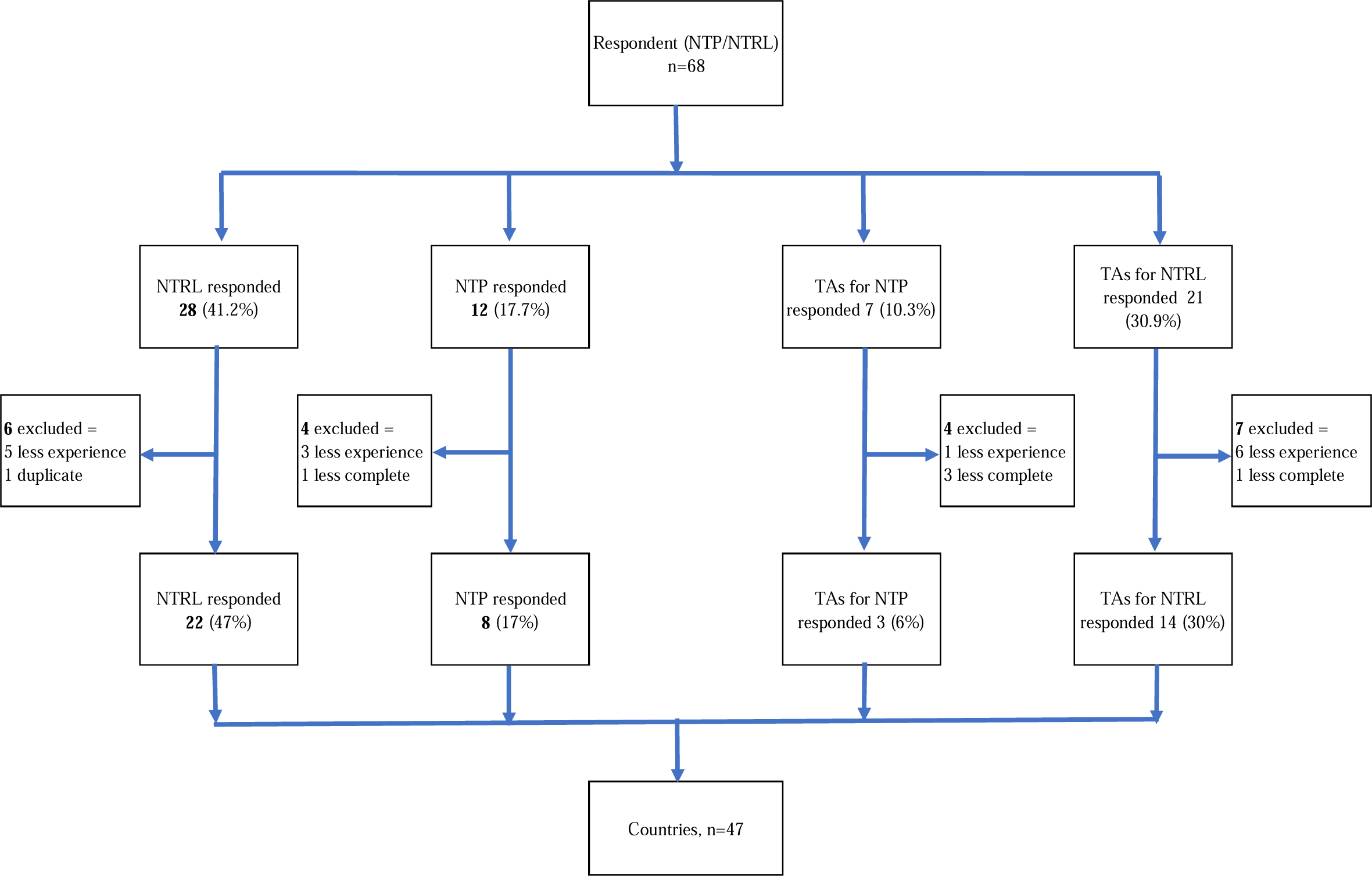
Number and category of responders. Key: NTRL managers, National TB Reference Laboratory managers; NTP managers, National TB Programme managers and TAs, Technical Assistants for NTRL and NTP.

### Laboratory Governance

Of the 47 country respondents, 25 (53%) experienced national security issues between 2007 and 2021 that affected in-country health services (e.g.: civil wars, strikes, health emergencies like COVID-19, Ebola, Cholera, etc..), 34 (72%) had national laboratory policy in place, and 32 (68%) had DLS within the MoH of which 32 (56%) were established between 2006-2021. A total of 31 countries (66%) had National TB Laboratory Strategic Plans (LSP) of which 18 (58%) are integrated into the National TB strategic plans. Only 14 (45%) LSP have a comprehensive annual budget line. Of 47 countries, 44 (94%) have the NTRLs formally recognized by the MoH of which 27 (57%) are classified as TB-containment Laboratories (high-risk TB Laboratories or Biosafety Level 3). When it comes to the influencers in the uptake of TB diagnostic technologies at the country level, of the 47 respondents, 32 (68%) ranked Political Commitment as the highest followed by recommendations raised from in-country technical assistance mission reports 24 (51%), (**Table 1**). The MoH was ranked the highest as the funder of the NTRL operationalization and a driver of the adoption, adaptation, and implementation of new TB diagnostic technologies with 18 (38%) and 27 (57%) respondents respectively (**Figure S2. a and b**) Considering and combining the MoH, DLS, and NTP as domestic (national) and Partners with WHO (HQ, AFRO and Country Offices) together as international, on average, 36% had no response while 35% of respondents stated the source of funds for NTRL were domestic. As the driver of the whole implementation, the combination showed us that 52% of respondents ranked domestic (national=MoH, DLS, NTP, and NTRL) as the highest driver/funding source.

**Table 1.**
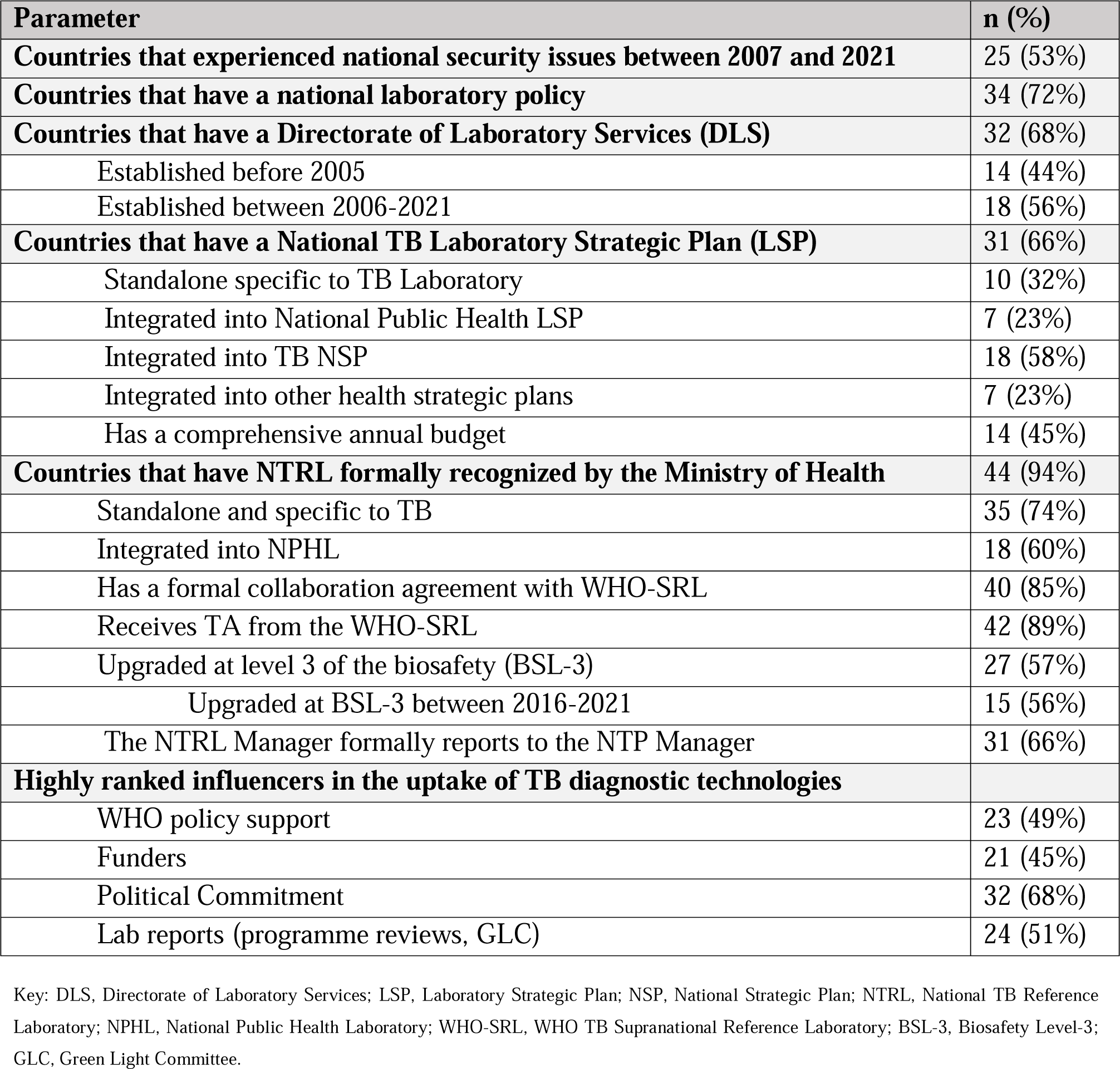
The characteristics of respondent countries: Laboratory Governance.

| **Parameter** | **n (%)** |
| --- | --- |
| **Countries that experienced national security issues between 2007 and 2021** | 25 (53%) |
| **Countries that have a national laboratory policy** | 34 (72%) |
| **Countries that have a Directorate of Laboratory Services (DLS)** | 32 (68%) |
| Established before 2005 | 14 (44%) |
| Established between 2006-2021 | 18 (56%) |
| **Countries that have a National TB Laboratory Strategic Plan (LSP)** | 31 (66%) |
| Standalone specific to TB Laboratory | 10 (32%) |
| Integrated into National Public Health LSP | 7 (23%) |
| Integrated into TB NSP | 18 (58%) |
| Integrated into other health strategic plans | 7 (23%) |
| Has a comprehensive annual budget | 14 (45%) |
| **Countries that have NTRL formally recognized by the Ministry of Health** | 44 (94%) |
| Standalone and specific to TB | 35 (74%) |
| Integrated into NPHL | 18 (60%) |
| Has a formal collaboration agreement with WHO-SRL | 40 (85%) |
| Receives TA from the WHO-SRL | 42 (89%) |
| Upgraded at level 3 of the biosafety (BSL-3) | 27 (57%) |
| Upgraded at BSL-3 between 2016-2021 | 15 (56%) |
| The NTRL Manager formally reports to the NTP Manager | 31 (66%) |
| **Highly ranked influencers in the uptake of TB diagnostic technologies** |  |
| WHO policy support | 23 (49%) |
| Funders | 21 (45%) |
| Political Commitment | 32 (68%) |
| Lab reports (programme reviews, GLC) | 24 (51%) |

Key: DLS, Directorate of Laboratory Services; LSP, Laboratory Strategic Plan; NSP, National Strategic Plan; NTRL, National TB Reference Laboratory; NPHL, National Public Health Laboratory; WHO-SRL, WHO TB Supranational Reference Laboratory; BSL-3, Biosafety Level-3; GLC, Green Light Committee.

### Policy endorsement and implementation status for TB Diagnostics

The implementation of TB diagnostic technologies after the endorsement by WHO has been influenced by many factors based on the laboratory preparedness, technology transfer, and routine testing phases in all 47 countries in the WHO/AFR that responded to the survey and showed that it took from two to nine years from the time the test is endorsed until it is fully used at the routine testing phase (**Table 2**)

**Table 2.** Policy endorsement and implementation status for TB diagnostics.

| **Parameter** | **n (%)** |
| --- | --- |
| **MGIT (Liquid culture & speciation), 2007** |  |
| Laboratory preparedness (2007-2011) | 22 (47%) |
| Technology transfer (2012-2015) | 21 (45%) |
| Routine testing ($\geq 2016$) | 30 (64%) |
| **LPA (first and second-line), 2008** |  |
| Laboratory preparedness (2008-2011) | 19 (40%) |
| Technology transfer (2012-2015) | 25 (53%) |
| Routine testing ($\geq 2016$) | 34 (72%) |
| **DST (second-line phenotypic), 2008** |  |
| Laboratory preparedness (2008-2011) | 15 (32%) |
| Technology transfer (2012-2015) | 21 (45%) |
| Routine testing ($\geq 2016$) | 31 (66%) |
| **LED Microscopy, 2010** |  |
| Laboratory preparedness (2010-2013) | 26 (55%) |
| Technology transfer (2014-2017) | 35 (74%) |
| Routine testing ($\geq 2017$) | 37 (79%) |
| **Non-Commercial Culture and DST, 2010** |  |
| Implemented | 5 (11%) |
| *CRI (Colorimetric Redox Indicator) method in use for Culture and DST* | 4 (9%) |
| *NRA (Nitrate Reductase Assay) method in use for Culture and DST* | 2 (4%) |
| *MODS (Microscopic Observation Drug Susceptibility Assay) method in use for Culture and DST* | 3 (6%) |
| Laboratory preparedness (2010-2013) | 4 (9%) |
| Technology transfer (2014-2017) | 5 (11%) |
| Routine testing ($\geq 2017$) | 5 (11%) |
| **Xpert MTB/RIF, 2010** |  |
| All adult and children suspected of TB | 39 (83%) |
| Laboratory preparedness (2010-2013) | 32 (68%) |
| Technology transfer (2014-2017) | 40 (85%) |
| Routine testing ($\geq 2017$) | 43 (91%) |
| **LF-LAM, 2015** |  |
| Implemented | 23 (49%) |
| Used to test TB in PLHIV seriously ill only | 19 (40%) |
| Test used for all adults and children | 19 (40%) |
| Used in limited number of hospitals | 7 (15%) |
| Laboratory preparedness (2015-2018) | 13 (28%) |
| Technology transfer (2018-2020) | 15 (32%) |
| Routine testing ($\geq 2021$) | 22 (47%) |
| **TB LAMP, 2016** |  |
| Implemented | 13 (28%) |
| Laboratory preparedness (2015-2018) | 4 (9%) |
| Technology transfer (2019-2020) | 9 (19%) |
| Routine testing ($\geq 2021$) | 10 (21%) |
| **Xpert ULTRA, 2017** |  |
| Implemented (transition from Xpert MTB/RIF to Xpert ULTRA) | 44 (94%) |
| Laboratory preparedness (2017-2020) | 28 (60%) |
| Technology transfer (2019-2020) | 42 (89%) |
| Routine testing ($\geq 2021$) | 34 (72%) |
| **TrueNat TB/RIF, 2019** |  |
| Implemented | 12 (26%) |
| Laboratory preparedness (2019-2020) | 4 (9%) |
| Technology transfer ($\geq 2021$) | 9 (19%) |
| Routine testing ($\geq 2021$) | 7 (15%) |
| **Xpert MTB/XDR, 2019** |  |
| Laboratory preparedness (2019-2020) | 5 (11%) |
| Technology transfer ($\geq 2021$) | 27 (57%) |
| Routine testing ($\geq 2021$) | 20 (43%) |

Key: MGIT, Mycobacterium Growth Indicator Tube; LPA, Line Probe Assay; DST, Drug Susceptibility Testing; LED, Light-Emitting Diodes; CRI, Colorimetric Redox Indicator; NRA, Nitrate Reductase Assay; MODS, Microscopic Observation Drug Susceptibility; Xpert MTB/RIF, Xpert Mycobacterium tuberculosis/Rifampicin; LF-LAM, Lateral Flow Lipoarabinomannan; TB LAMP, TB Loop-Mediated Isothermal Amplification; Xpert ULTRA, Xpert MTB/RIF ULTRA; Xpert MTB/XDR, Xpert Mycobacterium tuberculosis/ Extensively drug-resistant TB.

MGIT liquid culture and speciation – 1^st^ line LPA and 2^nd^ line phenotypic DST endorsed by 2007-2008, and by 2016 (1^st^ & 2^nd^ line LPA), of the 47 countries, 30 (64%); 34 (72%), and 31 (66%) were in the routine testing phase for MGIT (liquid culture & speciation); LPA (first and second-line), and DST (second-line phenotypic) respectively. Relatedly, LED Microscopy, rapid non-Commercial culture, and DST, and Xpert MTB/RIF which were endorsed in 2010, at the time of the survey, almost 5 (11%), and 39 (83%) countries had adopted and implemented these policies respectively. By 2017, almost 37 (79%) for LED microscopy, 5 (11%) for non-commercial culture and DST methods, and 43 (91%) for Xpert MTB/RIF were in the routine testing phase.

Reviewing the status of the uptake of LF-LAM endorsed in 2015 and TB-LAMP endorsed in 2016, at the time of the survey, 23 (49%) respondents confirmed to have adopted LF-LAM and 13 (28%) TB-LAMP respectively as TB diagnostic test in their countries. As of 2021, almost 22 (47%) for LF-LAM and 10 (21%) for TB LAMP were in the routine testing phase.

Considering the Xpert ULTRA endorsed in 2017, TrueNat MTB/RIF and Xpert MTB/XDR endorsed in 2019, a total of 44 (94%) have adopted and started the implementation for Xpert ULTRA, and 12 (26%) for TrueNat MTB/RIF respectively and as of 2021, 34 (72%) for Xpert ULTRA, 7 (15%) for TrueNat TB/RIF, and 20 (43%) for Xpert MTB/XDR were in the routine testing phase.

Our study showed us that the uptake of technologies varied from country to country and was driven by the burden of TB, TB/HIV, and MDR-TB in Africa hence the majority of countries classified as rapid implementers are in the East and Southern Africa subregion (where the burden of diseases is high) while others were moderate and even slow implementers in the Central and Western Africa subregion (**Figure 2**)

**Figure 2.**
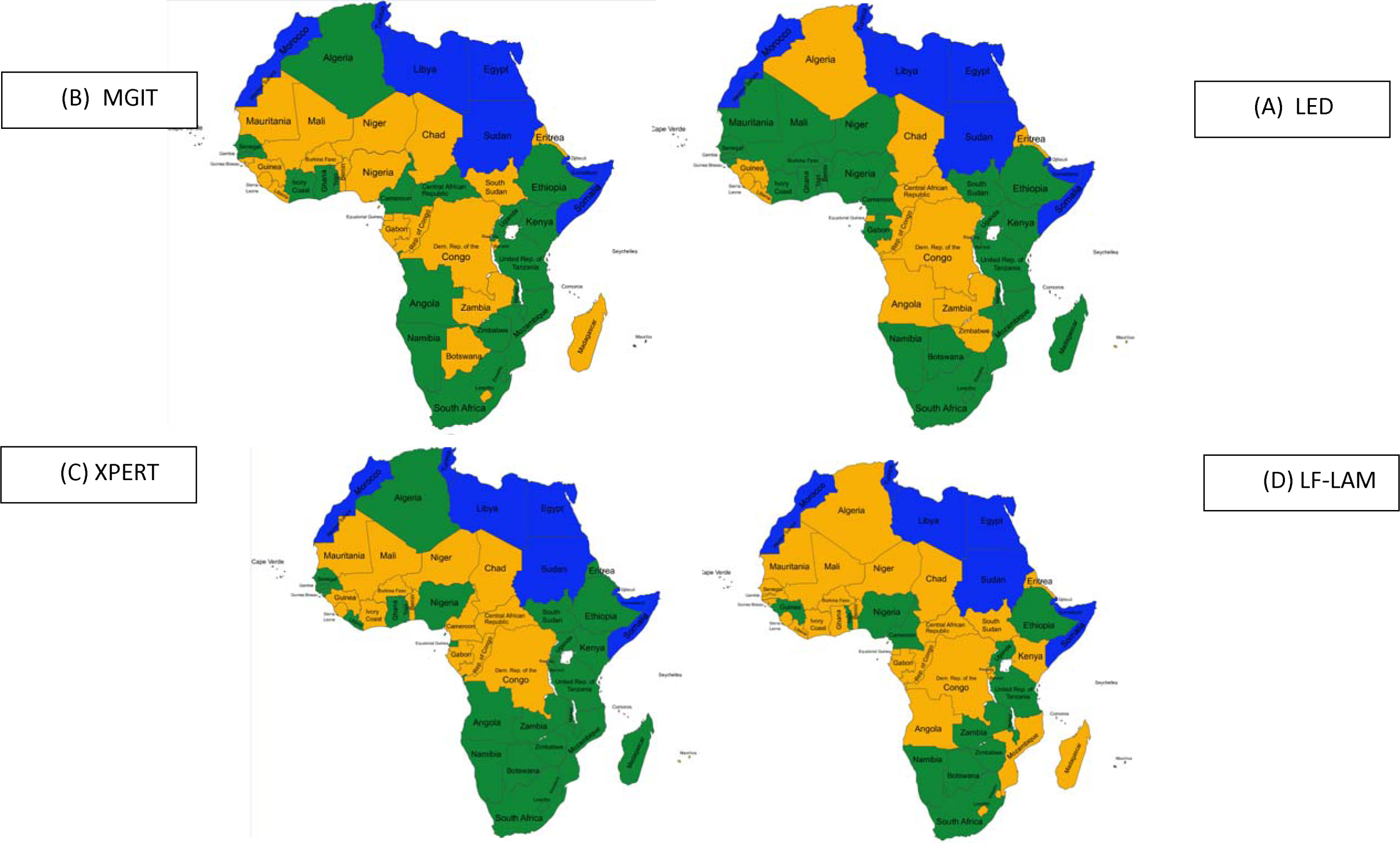
Rapid, moderate, and slow implementers of (A) MGIT, (B) LED, (C) XPERT, and (D) LF-LAM in WHO/AFR. Key: MGIT, Mycobacterium Growth Indicator Tube; LED, Light Emitting Diodes Fluorescence microscopy; LF-LAM, Lateral Flow Lipoarabinomannan assay. Blue color: countries outside of WHO/AFR; Orange color, countries that are moderate/slow implementers (within 3 to 5 years/5 years onward after endorsement); green color, countries that are rapid implementers ( within 1-3 years).

### Predisposing, enabling, and need factors influencing the uptake of WHO-endorsed TB diagnostic technologies

PEN factors influencing the uptake of TB diagnostic technologies are summarized with specific tests (**Table 3**). The predisposing including, competent staff and lab preparedness are ranked first and second key factors, respectively. The enabling factors that ranked highly were the availability of funds, political commitment, and SRL support. Relatedly, the need factors that ranked highest were the increase in TB incidence and mortality as well as the emergency of MDR-TB, especially for MGIT, 2^nd^ line phenotypic DST, and LPA.

**Table 3.**
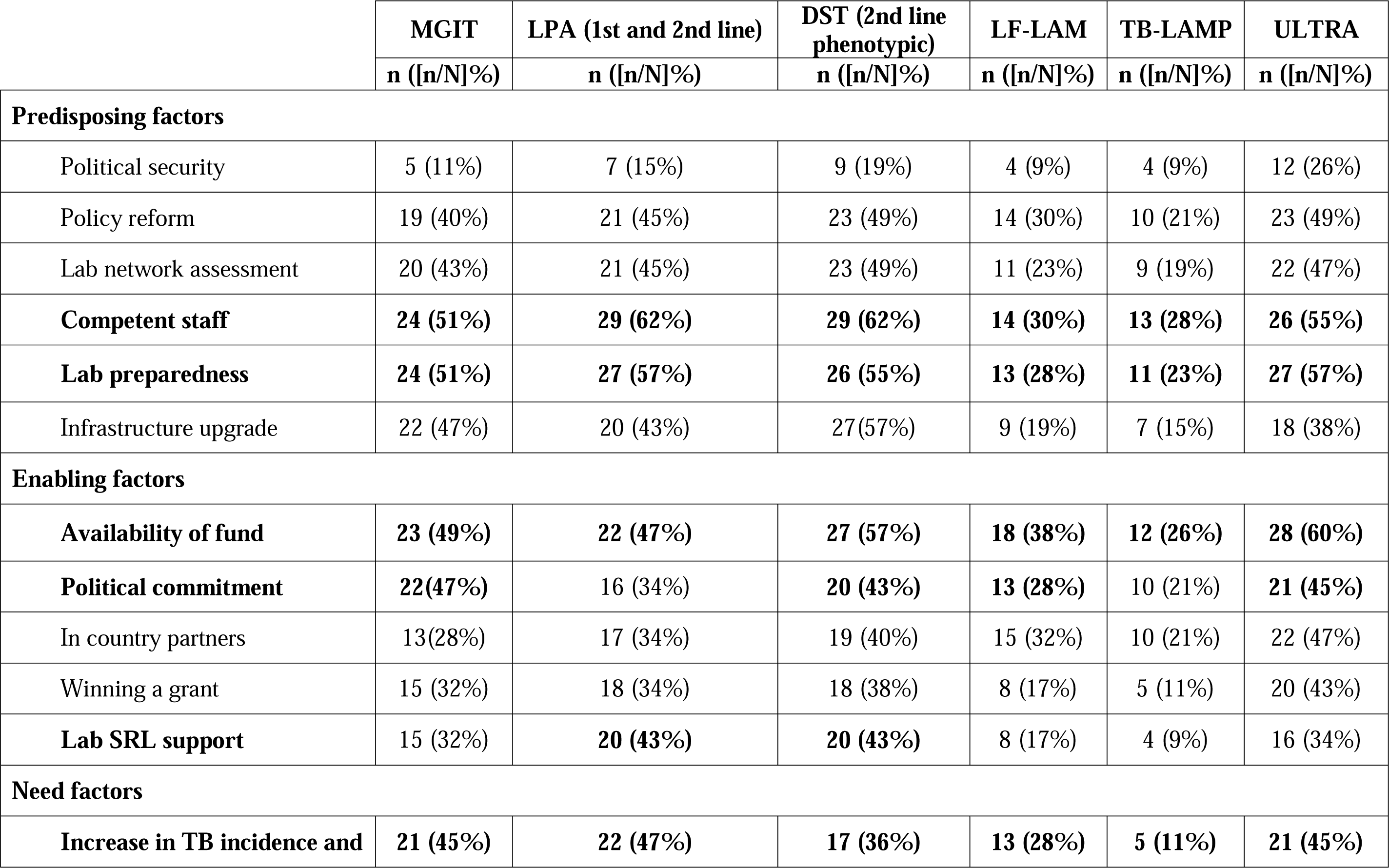
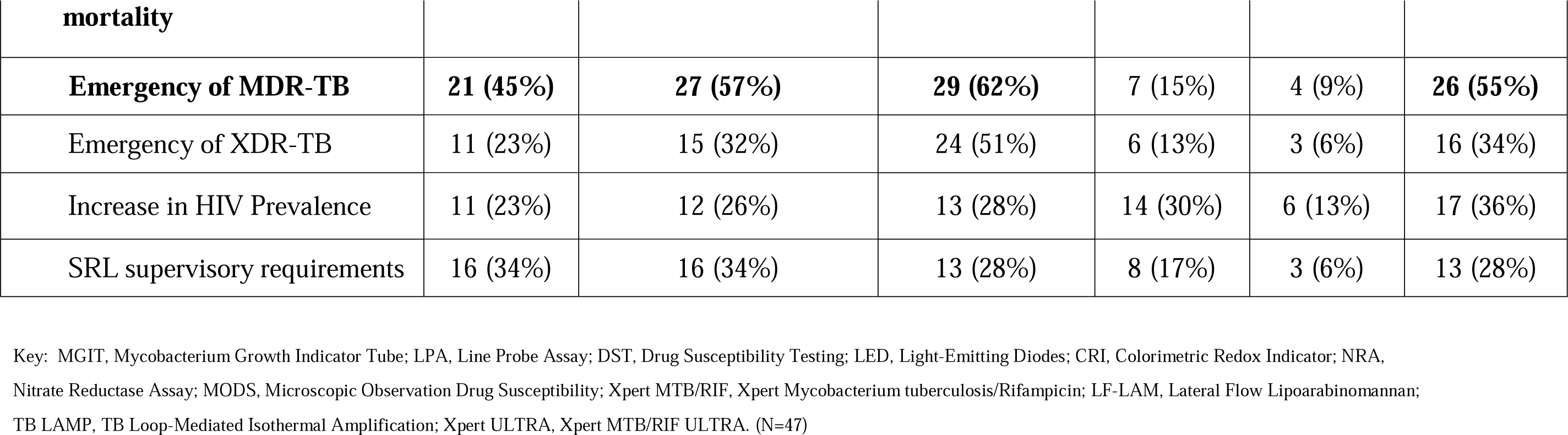
Predisposing, Enabling, and Need factors influencing the uptake of TB diagnostic technologies.

|  | **MGIT** | **LPA (1st and 2nd line)** | **DST (2nd line phenotypic)** | **LF-LAM** | **TB-LAMP** | **ULTRA** |
| --- | --- | --- | --- | --- | --- | --- |
|  | **n ([n/N]%)** | **n ([n/N]%)** | **n ([n/N]%)** | **n ([n/N]%)** | **n ([n/N]%)** | **n ([n/N]%)** |
| **Predisposing factors** |  |  |  |  |  |  |
| Political security | 5 (11%) | 7 (15%) | 9 (19%) | 4 (9%) | 4 (9%) | 12 (26%) |
| Policy reform | 19 (40%) | 21 (45%) | 23 (49%) | 14 (30%) | 10 (21%) | 23 (49%) |
| Lab network assessment | 20 (43%) | 21 (45%) | 23 (49%) | 11 (23%) | 9 (19%) | 22 (47%) |
| **Competent staff** | **24 (51%)** | **29 (62%)** | **29 (62%)** | **14 (30%)** | **13 (28%)** | **26 (55%)** |
| **Lab preparedness** | **24 (51%)** | **27 (57%)** | **26 (55%)** | **13 (28%)** | **11 (23%)** | **27 (57%)** |
| Infrastructure upgrade | 22 (47%) | 20 (43%) | 27(57%) | 9 (19%) | 7 (15%) | 18 (38%) |
| **Enabling factors** |  |  |  |  |  |  |
| **Availability of fund** | **23 (49%)** | **22 (47%)** | **27 (57%)** | **18 (38%)** | **12 (26%)** | **28 (60%)** |
| **Political commitment** | **22(47%)** | 16 (34%) | **20 (43%)** | **13 (28%)** | 10 (21%) | **21 (45%)** |
| In country partners | 13(28%) | 17 (34%) | 19 (40%) | 15 (32%) | 10 (21%) | 22 (47%) |
| Winning a grant | 15 (32%) | 18 (34%) | 18 (38%) | 8 (17%) | 5 (11%) | 20 (43%) |
| **Lab SRL support** | 15 (32%) | **20 (43%)** | **20 (43%)** | 8 (17%) | 4 (9%) | 16 (34%) |
| **Need factors** |  |  |  |  |  |  |
| **Increase in TB incidence and** | **21 (45%)** | **22 (47%)** | **17 (36%)** | **13 (28%)** | **5 (11%)** | **21 (45%)** |

| mortality |  |  |  |  |  |  |
| --- | --- | --- | --- | --- | --- | --- |
| **Emergency of MDR-TB** | **21 (45%)** | **27 (57%)** | **29 (62%)** | 7 (15%) | 4 (9%) | **26 (55%)** |
| Emergency of XDR-TB | 11 (23%) | 15 (32%) | 24 (51%) | 6 (13%) | 3 (6%) | 16 (34%) |
| Increase in HIV Prevalence | 11 (23%) | 12 (26%) | 13 (28%) | 14 (30%) | 6 (13%) | 17 (36%) |
| SRL supervisory requirements | 16 (34%) | 16 (34%) | 13 (28%) | 8 (17%) | 3 (6%) | 13 (28%) |

Key: MGIT, Mycobacterium Growth Indicator Tube; LPA, Line Probe Assay; DST, Drug Susceptibility Testing; LED, Light-Emitting Diodes; CRI, Colorimetric Redox Indicator; NRA, Nitrate Reductase Assay; MODS, Microscopic Observation Drug Susceptibility; Xpert MTB/RIF, Xpert Mycobacterium tuberculosis/Rifampicin; LF-LAM, Lateral Flow Lipoarabinomannan; TB LAMP, TB Loop-Mediated Isothermal Amplification; Xpert ULTRA, Xpert MTB/RIF ULTRA. (N=47)

### Impediments and barriers to rapid implementation of TB diagnostic technologies in countries

All 47 have reported impediments and barriers to implementing WHO-endorsed diagnostic technologies. A total of 33 (70%) ranked insufficient finance to procure the test as the highest followed by the procurement issues with 26 (55%) of respondents (**Figure S3**).

## DISCUSSION

This is the first survey to assess the status of the uptake and the PEN factors influencing the uptake of TB diagnostic technologies in the WHO African Region. Our survey showed that the uptake is slow and takes several years between the time a technology is endorsed to when it reaches the routine testing phase. East and Southern African countries are rapid implementers in general and across all diagnostic technologies, while in Central and West Africa, many countries are still lagging and slower, and extra effort is needed. There is also an unbalance in terms of funding and partner support to countries in East and Southern Africa versus Central and Western Africa. The uptake requires in-country ownership with a well-established structure at the level of the Ministry of Health to drive the laboratory system and address TB diagnostic needs. This is confirmed by 32 (68%) respondents who recognized political commitment as a highly influencing factor for the TB diagnostic uptake and our finding is in agreement with the statement of the Lancet Commission on diagnostics by Fleming et al.^26^ This also complements the findings from Pai et al., stating that the availability of new tools does not mean that they will be adopted, used correctly, scaled up or have public health impact.^27^ Human resources for the laboratory, availability of funds and increase in TB incidence including mortality are predisposing, enabling, and need factors, respectively, influencing TB diagnostic uptake. Almost 72% of countries have national laboratory policy in place, 45% increase from the 26.5% reported by Ondoa et al. in 2017. With 31 (66%) countries that have National Laboratory Strategic plans specific to TB, our study shows more than twice an increase from the 28.6% we previously reported in 2017.^28^ This is great progress in the African Region and it is in alignment with the 2008 Maputo declaration and the 2008 Regional Committee Resolution of the WHO Regional Office for Africa (WHO AFRO), ^29^ calling member states to establish national laboratory strategies and policies in a way to strengthen laboratory systems as an integral part of disease control ^30^.

In addressing TB specifically, 44 (94%) countries have NTRLs established and formally recognized by the MoH fulfilling their role in strengthening TB laboratory network capacity and 85% of them have a formal collaboration agreement with the WHO-SRL network showing a small increase compared to the number from our previous study that reported 81.8% of NTRL that had collaboration agreements ^28^. As result of this collaboration, the WHO-SRL Uganda through its technical assistance to 21 countries in Africa, contributed to the increase from two in 2015 to 8 NTRLs ISO 15189:2012 accredited by 2022,^31^ and more laboratories are achieving this milestone. This demonstrates the critical role the SRL network is playing in mentoring the NTRLs through regional-driven laboratory projects to technically assist countries in strengthening the quality of TB laboratory services. Through different phases of laboratory preparedness, technology transfer, and routine testing, our study showed that the impact of a technology can be measured between two and 9 years after the endorsement. For instance, the uptake of MGIT (Liquid culture & speciation) and LPA (1^st^ &2^nd^ line) after their endorsement in 2007 and 2008 respectively, took over 8 years for LPA and 9 years for MGIT to reach the routine testing phase in 2016, and with only 64% and 72% of the countries reporting laboratory results using those tests respectively. The delay in their uptake as shown by our findings is similar to that reported by Maningi et al.,^8^ and also this confirms data from the recent study in the WHO European Region where two thirds of the NTRLs were able to perform DST for Bedaquilin (BDQ) over 9 years after its endorsement by WHO.^32^ Both MGIT and LPA have the advantage of being rapid and highly sensitive in diagnosing TB, however, they require high skills and specialized infrastructure such a containment laboratory and the 3-rooms, respectively. It is clear that the laboratory preparedness phase during which the infrastructure, among other requirements, has to be fixed before the technology could be transferred, took much time and influenced this delay.

By 2017, almost seven years after LED Fluorescence Microscopy (LED FM) endorsement, 79% of countries were at the stage of using it routinely and this is in line with other findings stating that LED fluorescence microscopy gives a legitimate option in contrast to conventional ZN techniques in terms of its higher sensitivity, time-saving, and minimal effort.^33^

The implementation of Xpert MTB/RIF after 2010 and later 2013 shows that it has been optimal in 41 (91%) countries which were at the routine testing phase, 7 years since its first endorsement in 2010. At the time of the survey, 39 (83%) confirmed to have adopted the policy and started the implementation. By 2013, within 4 years after the additional recommendation, almost 32 (68%) countries were in the laboratory preparedness phase which shows how Xpert has been considered a breakthrough technology in diagnosing TB and RIF-resistant TB. This uptake was quicker and well accepted than LPA for instance where 19 (40%) were in the laboratory preparedness phase by 2011, almost 3 years after LPA endorsement. Besides that, Xpert faced health system challenges related to modular failures, poor power supply, inefficient sample transport mechanisms, weak data management, and inadequate human resources to staff the remote test sites which did not help increase TB case notification in Nigeria, DRC and South Africa as demonstrated by Williams et al,. ^34^ After 2013 with additional recommendations on the use of Xpert for children and extrapulmonary tuberculosis, an improvement in its use was noticed with 40 (85%) countries moving to the technology transfer phase between 2014 and 2017. Then Xpert Ultra (Ultra) came on board in 2017, and as of 2021, almost 34 (72%) countries were in the routine testing phase using Ultra showing that the laboratory preparedness phase did not require extra efforts at the country level, instead, countries had to transition from Xpert MTB/RIF use to Ultra using the same infrastructure. The awareness of the use of Xpert as a platform contributed positively at the time of the survey, 44 (94%) countries confirmed starting the implementation of Ultra. However, the country’s transition was slow. The transition to Ultra was motivated by the fact that Cepheid stopped manufacturing the Xpert MTB/RIF cartridges.

LF-LAM policy was adopted and implemented in 23 (49%) countries at the time of the survey. By 2021, over 5 years after the establishment of the WHO policy on LF-LAM, our survey shows that 22 (47%) countries were routinely using LF-LAM. Our results are similar to what Singhroy et al. found while conducting a survey in high HIV burden countries which was 46% of countries implementing LF-LAM ^35^.

In 2016, the TB-LAMP policy was published by WHO, and at the time of the survey, only 13 (28%) countries had adopted the policy. By 2021, over 5 years after its recommendation, only 10 (21%) were routinely reporting data using TB-LAMP. The fact that the technology does not provide a resistance profile might have limited its implementation at the country level. This may have been compounded by the rapid uptake of the available better alternative, GeneXpert being available at the same time for countries to choose from. Also, having 22 out of 47 as high HIV/TB burden countries in Africa may have limited its use, where Xpert is better recommended.^36^

In 2019, TrueNat TB/RIF was endorsed and at the time of the survey, the policy was adopted in 12 (26%) countries. By 2021, over 2 years after its endorsement, 7 (15%) countries were routinely using TrueNat and reporting data. The country’s willingness to include it in their algorithm is progressing, and also the fact it was endorsed at the time countries finished their plans and budgets and have already applied to the Global Fund for funding requests, was a limitation factor for its quick implementation. As of 2021, almost 20 (43%) countries were in the routine testing phase for the use of Xpert MTB/XDR. This quick uptake, a few years after its recommendation (2 years) could be linked to the fact that no additional training was needed for technicians who had previous experience using Xpert Ultra or a minimum training, of at least one day was needed for those without previous experience with. Our findings are in line with those reported recently by Katamba et al. ^18^

As a funder of the NTRL and driver of the implementation, the Ministry of Health is ranked highest for both, with 18 (38%) and 27 (57%) respondents respectively. On average, 35% of respondents found that funding for the NTRL is from domestic and the driver of the TB diagnostics implementation is national for 52% of respondents. The 2023 Global TB Report data^3^ tells us that funding for TB in 2022, around 60% was international (**Figure S4**). In comparison with our findings and knowing that funds for the lab are international, respondents may not be aware of the laboratory budget and source, or may not be fully involved in the budget plans for the laboratory. This may be in line and confirm our findings with 14/31 (45%) of respondents who stated that the annual budget of the National TB Laboratory Strategic plan was in place, which is a small number (**Table 1**)

### Predisposing, Enabling, and Need factors influencing the uptake of diagnostic technologies

**Predisposing factors:** the competence of staff and laboratory preparedness were highly ranked and varied across all the technologies. Regarding the competence of staff, our findings are comparable to those highlighted in the Lancet Commission paper on diagnostics ^26^ stating that technological innovation goes along with workforce empowerment and capacity building. Our survey shows that out of 47 countries, 25 (53%) experienced national security issues between 2007 and 2021 (e.g. political unrest, strike, disease outbreaks like Ebola, Covid-14, etc.) that affected the health system. This is comparable to findings from Cote d’Ivoire (CI) which were reported by Ekaza et al. who stated that military and political conflict in CI favored the spread of infectious diseases affecting the workforce, TB being among the most devastating ^37^.

On the laboratory preparedness side, our data show that the preparedness phase, after any technology endorsement varies from country and is based on the laboratory requirement per technology. This preparedness phase was given 3 years as duration which involved policy reform, laboratory assessment, infrastructure upgrade, and creation of the Standard Operating Procedures. Xpert MTB/RIF highly ranked first with 68%, then Ultra with 60%, and LED Microscopy with 55% of countries being in the preparatory phase three years after endorsement. Our findings confirm that the majority of countries started their laboratory preparatory phases once those technologies were endorsed, however, they were delayed in getting into the routine testing phase and some requirements needed to be fixed before the technologies could be transferred to countries such as training, validation of the technology, quality assurance, procurement process, etc., which may have been the cause. For instance, once the policy on the use of Xpert was issued in 2010, WHO developed a manual on the “know-how to” implement to assist countries in being prepared before they uptake this technology ^38^. MGIT (liquid culture and speciation) and LPAs faced the issues of the infrastructure requirement.

**Enabling factors:** The availability of funds is ranked first for all the technologies and Political commitment, in-country partners (e.g.: for LF-LAM as point of care and Ultra) and TB Supranational Reference Laboratory support (for LPAs and 2^nd^ line phenotypic DST) second for some others. Technical assistance was important to transfer knowledge where partners and SRL play an important role in the implementation.

**Need factors:** the emergency of MDR-TB was highly ranked for 2^nd^ line phenotypic DST with 62%, for LPA (1^st^ and 2^nd^ line) with 57% and for Ultra with 55%. Another factor was the increase in TB incidence and mortality for LPA (1^st^ and 2^nd^ line) with 57% first, followed by MGIT (liquid culture and speciation) and Ultra both with 45% and 2^nd^ line phenotypic DST with 36% of respondents.

As limitations, it is apparent that the uptake of diagnostic technologies has been driven by donor funding and priorities for disease burden. Most of the donors do not fund all 47 countries and all the countries are not implementing technologies equally. Therefore, some countries (low TB, TB/HIV, and MDR-TB burden) showed a delay in the uptake, suffered from some barriers and some of them have no DLS nor formal NTRL, no LSP to address TB laboratory requirements, etc.. For some countries, there was a delay in getting feedback from respondents as a consequence of poor response rate on a survey. The lack of experience of some respondents affected also a part of the quality and completeness of the survey

As strengths of the study, almost 36 (77%) respondents were from the NTRL Managers and technical assistants for the NTRL. Their responses were consistent in addressing laboratory data concerns as being at the frontline for uptake and well-positioned for implementing WHO-TB diagnostic technologies as they become available. Having also NTP Manager’s response completed missing information if any from the other respondents was a strength. All the 47 countries in WHO/AFR responded and our analysis covered all the countries.

As a recommendation on the required efforts for speeding up the uptake, the implementation guide or tools should be available for each diagnostic technology upon its endorsement.

## CONCLUSION

Besides the availability of diagnostic technologies and the existence of the WHO Guidelines for their implementations, factors influencing their uptake to improve access at the country level are multiple. It can take from two to nine years before a new TB diagnostic test can go to the routine testing phase after endorsement. The political commitment to address laboratory policy, the establishment of a Directorate of Laboratory Services with a formal recognition of the National TB Reference Laboratory within the Ministry of Health and the existence of the budgeted laboratory strategic plans are key. After that, workforce empowerment, laboratory preparedness to improve infrastructure capacity, in-country partnership to map the technical assistance, the support of the TB Supranational Reference Laboratory network, and the incidence, mortality, and emergency of MDR-TB could be taken into account as predisposing, enabling and needs factors influencing the uptake of TB diagnostic technologies for the TB control programme. Every diagnostic technology, upon endorsement by WHO, should be accompanied by the “know-how to” implement guidance and funding for its rapid uptake and roll out. it is recommended to advocate for more resources to strengthen TB laboratory capacity in Central and West Africa along with maintaining the gains from the East and Southern African Regions where the burden of TB, TB/HIV, and MDR-TB is still high.

## Supporting information

Map of 47 participant countries

Ranked funders and drivers

Ranked impediments and Barriers

Funding for TB in Africa

Supplementary file 1-Survey questionnaire

## Data Availability

All relevant data are within the paper and its supporting information file

https://docs.google.com/forms/u/0/

## Acknowledgments

We would like to thank the National TB Programme (NTP) and National TB Reference Laboratory (NTRL) managers, key partners, and officials from the Ministry of Health in the 47 Member States for responding to the survey as well as the WHO country office and SRL Uganda representatives for their assistance in disseminating the survey. Special thanks go to Mr. Shaffic Ssettimba for assisting in designing the survey.

## Declarations

the first author works for the World Health Organization (WHO). The contents of this article are the author’s personal opinion.

## Availability of Data and Materials

All relevant data are within the paper and its supporting information file1

## Conflict of interest

None declared

## Notes

### Competing Interest Statement

The authors have declared no competing interest.

### Clinical Protocols

https://www.isrctn.com/ISRCTN24711056

### Funding Statement

This study did not receive any funding.

### Author Declarations

The questionnaire was submitted together with the study protocol to the Research and Ethics Committee of the School of Biomedical Sciences (SBS-2022-154) at the College of Health Sciences, Makerere University, Kampala, Uganda, and to the Uganda National Council for Science and Technology (HS2393ES) for review and approval before it is converted to an online survey.

