## Supplementary material for "Predisposing, enabling, and need factors influencing rapid uptake of the World Health Organization-endorsed TB diagnostic technologies in Africa": Map of 47 participant countries

### Slide 1
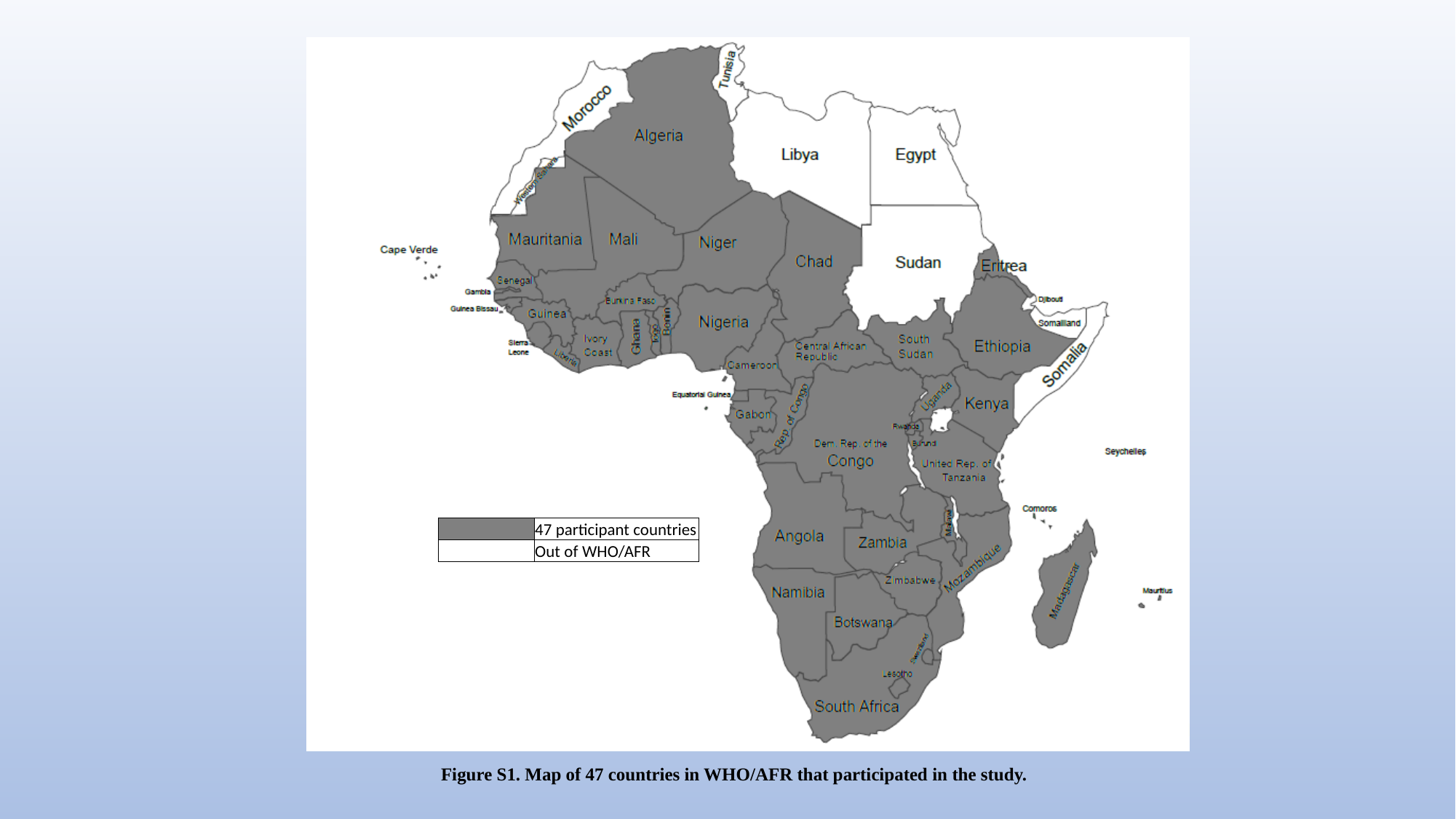

| | 47 participant countries |
| --- | --- |
| | Out of WHO/AFR |
#### Figure S1. Map of 47 countries in WHO/AFR that participated in the study.
