## Supplementary file 1-Survey questionnaire for "Predisposing, enabling, and need factors influencing rapid uptake of the World Health Organization-endorsed TB diagnostic technologies in Africa"

Project: predisposing, enabling and need factors influencing rapid uptake of WHO-endorsed TB diagnostic technologies in Africa

Principal Investigator: Jean de Dieu IRAGENA

Dear Colleague,

Apologies if you have received this message before and/or if you have already responded to it and/or if you get it multiple times. I have made use of contact lists which may overlap.

On top of my work, I am a PhD candidate at Makerere University College of Health Sciences, School of Biomedical Sciences, Kampala, Uganda. My research topic is ”***Roll-out and Scale-up of WHO-endorsed technologies for TB diagnosis in Africa: opportunities, strengths and challenges***” . I am now at the data collection stage and I am soliciting responses for my online survey from the TB laboratory managers, TB Programme managers, technicians, other officials from Ministries of Health (MoHs) and their partners.

I am counting on colleagues like you to achieve my study objective. Data from this survey will be used to assist countries in a better planning position to address factors that influence the uptake of WHO-endorsed diagnostic technologies for TB as they become available. I would therefore really appreciate it if you could spend 25 to 30 minutes to respond to my online survey, if you haven’t already done so.

Please use the link below (and send it on to your other colleagues if possible) to confirm your consent and to complete the survey. I would be very grateful for your support.

**INFORMED CONSENT SECTION**

Thank you for participating in this questionnaire survey “***predisposing, enabling and need factors influencing rapid uptake of WHO-endorsed TB diagnostic technologies***”. This survey is for academic purpose and it is conducted by Jean de Dieu IRAGENA (Principal Investigator) and Prof Moses JOLOBA, Dr Willy SSENGOOBA and Dr Achilles KATAMBA (Investigators Team) of the College of Health Sciences, School of Biomedical Sciences in Makerere University, Kampala, Uganda as part of the study team.

This survey should not take more than 30 minutes of your time and the following requirements must be met in order to participate

1. You may agree to participate
2. You should be TB laboratory manager, TB Programme manager, key partner, official from the Ministry of Health

The purpose of this survey is to better understand the factors that contribute to quick, moderate and slow uptake of TB diagnostic technologies in Africa after their endorsement by the World Health Organization (WHO). Slow, moderate and rapid implementers among member states including stakeholder awareness, predisposing and enabling factors such as funding related to the TB diagnostic uptake, access, utilization and needs will be identified. The focus will be on TB diagnostic technologies endorsed by the WHO between 2007 and 2021.

There are no immediate personal benefits from participating in this survey except you will gain a better understanding of how TB diagnostic technologies are considered for uptake at country level and the different factors that influence this uptake.

Confidentiality in connection with this survey will be ensured and generated data will be used for research purpose only. You may withdraw your consent at any time during the survey and refuse to answer any questions you do not want to answer.

In case of any question concerning this study, please contact Jean de Dieu IRAGENA (Principal Investigator) at +256783422722 or +242065117992 or

This survey questionnaire has received ethics clearance through the Research and Ethics Committee of the School of Biomedical Sciences (SBS-REC) at the College of Health Sciences, Makerere University, Kampala, Uganda. You can phone the School of Biomedical Sciences Research Ethics Committee at +256752575050 or if there is still something not explained well to you, or if you have a complaint.

If you wish to participate in this survey, please click the “I agree to participate” button below. If you do not wish to participate, please click the “I decline to participate” button below.

Do you consent to participate in this survey ?

☐ I agree to participate

☐ I decline to participate

I've invited you to fill out a form:

**DATA COLLECTION SECTION**

Questionnaire: predisposing, enabling and need factors influencing rapid uptake of WHO-endorsed TB diagnostic technologies

**Predisposing factors** (political security, policy reform, laboratory network assessment, competent staff, training, laboratory preparedness to uptake diagnostics, infrastructure upgrade, laboratory validation, quality assurance), **enabling factors** (availability of funding, political commitment, in country partners, winning grant, donation, procurement, etc.) as well as **needs** (emergence of drug resistant TB, increase of TB incidence) influencing WHO technology implementation will be emphasized and collected

**Targets:** TB laboratory managers, programme managers, technicians, other officials from Ministries of Health (MoHs) and their partners.

**Focus:** Predisposing, enabling factors and needs

Please fill in your contact information. This information will only be used to contact you about your survey response and will not be shared with any third parties.

**Name**

**Function**

**Email**

**Institution**

**Country**

Please select response that applies and specified when needed:

**Q1**: did your country experience **national security** that affected the implementation of TB diagnostic technologies between 2007 and 2021?

☐ Yes, if Yes, please select the appropriate period and explain what the issue was

| ☐ Between 1994 and 2005 | ☐ Between 2006 and 2015 | ☐ Between 2016 and 2021 |
| --- | --- | --- |
| Specify the exact year: | Specify the exact year: | Specify the exact year: |
| Explain what the issue was: | | |

☐ No

☐ I do not know

**Q2**: in your country, was there any **policy reform** related to laboratory system strengthening?

☐ Yes, if Yes, please select the appropriate period and what the reform was about

| ☐ Between 1994 and 2005 | ☐ Between 2006 and 2015 | ☐ Between 2016 and 2021 |
| --- | --- | --- |
| Specify the exact year: | Specify the exact year: | Specify the exact year: |
| Explain what the reform was about: | | |

☐ No

☐ I do not know

**Q3**: According to the **2008 Maputo declaration** on Strengthening of Laboratory Systems (available at <https://www.who.int/diagnostics_laboratory/Maputo-Declaration_2008.pdf>) , does your country have **National Laboratory Policy** within the national health development plan

☐ Yes, if Yes

☐ No

☐ I do not know

**Q4**: Does your country have a Laboratory Directorate (**Department of Laboratory Services)** within the Ministry of Health?

☐ Yes

| If Yes,please spell out the name: | | |
| --- | --- | --- |
| If Yes, please specify when it was established | | |
| ☐ Between 1994 and 2005 | ☐ Between 2006 and 2015 | ☐ Between 2016 and 2021 |
| ☐ Specify the exact year: | ☐ Specify the exact year: | ☐ Specify the exact year: |
| ☐ I do not know | | |

☐ No

☐ I do not know

**Q5**: does your country have a **National TB Reference Laboratory** (**NTRL**) formally recognized by the Ministry of Health?

☐ Yes

| If Yes, is your **National TB Reference Laboratory** | |
| --- | --- |
| **integrated** into the **National** **Public Health Laboratory?**  ☐ Yes  ☐ No  ☐ I do not know | a **standalone** TB laboratory only?  ☐ Yes  ☐ No  ☐ I do not know |

☐ No

☐ I do not know

**Q6**: does the **Head** of your National TB Reference Laboratory (NTRL) formally report to the **Head** of the National Tuberculosis Programme (NTP)?

☐ Yes

☐ No

If No, please specify the reporting line (to whom does he/she report?)

**Q7**: Does your country have **TB Laboratory Strategic Plan**?

☐ Yes

|  | If Yes, is your TB **Laboratory Strategic Plan** | |
| --- | --- | --- |
| **integrated** into the **National** **Public Health Laboratory Strategic Plan**?  ☐ Yes, if Yes, does it have a budget line?  ☐ Yes: what is the annual budget in USD?  ☐ No  ☐ I do not know  ☐ No  ☐ I do not know | **Integrated** into the **National TB Programme Strategic Plan?**  ☐ Yes, if Yes, does it have a budget line?  ☐ Yes: what is the annual budget in USD?  ☐ No  ☐ I do not know  ☐ No  ☐ I do not know | a **standalone** document specific to TB laboratory only?  ☐ Yes, if Yes, does it have a budget line?  ☐ Yes: what is the annual budget in USD?  ☐ No  ☐ I do not know  ☐ No  ☐ I do not know |

☐ No

☐ I do not know

**Q8**: does your NTRL have a **formal collaboration agreement** established within the WHO TB Supranational Reference Laboratory Network (SRLN)? See the list of the SRL <https://sites.google.com/site/srtblaboratories/>

If Yes, please spell out the name of the SRL:

If yes, please specify the nature of the support you receive from the SRL:

☐ Technical

☐ Financial

☐ Both: technical and financial

☐ No support at all

☐ Yes

| If Yes, specify the time when this collaboration agreement has been established | | |
| --- | --- | --- |
| ☐ Between 1994 and 2005 | ☐ Between 2006 and 2015 | ☐ Between 2016 and 2021 |
| ☐ Specify the exact year: | ☐ Specify the exact year: | ☐ Specify the exact year: |
| ☐ I do not know | | |

☐ No

☐ I d not know

**Q9**: at your country level, when a new diagnostic technology is endorsed by World Health Organization (WHO), who is driving the process of its **adoption**, a**daptation** and **implementation**?

☐ WHO Headquarters Geneva

☐ WHO Regional Office for Africa, Brazzaville

☐ WHO Country Office

☐ Ministry of Health

☐ Directorate of Laboratory Services

☐ National TB Programme

☐ National TB Reference Laboratory

☐ None of them

☐ All of them

Please select the **main body** that influence and motivates the uptake of technologies at your country level

☐ WHO

☐ Funders

☐ Government

☐ Laboratory reports and follow-up on action items from:

☐ TB Programme reviews

☐ Green Light Committee missions

☐ None of them

☐ All of them

**Q10**: Who is providing funds for the operationalization of your National TB Reference Laboratory activities?

☐ WHO Headquarters Geneva

☐ WHO Regional Office for Africa, Brazzaville

☐ WHO Country Office

☐ Ministry of Health

☐ Directorate of Laboratory Services

☐ National TB Programme

☐ Partners

☐ None of them

☐ All of them

**Q11**: did your country conduct a comprehensive **laboratory network assessment**?

☐ Yes

| If Yes, please specify when it was conducted | | |
| --- | --- | --- |
| ☐ Between 1994 and 2005 | ☐ Between 2006 and 2015 | ☐ Between 2016 and 2021 |
| ☐ Specify the exact year: | ☐ Specify the exact year: | ☐ Specify the exact year: |
| ☐ I do not know | | |

☐ No

☐ I do not know

**Q12**: does your country have in place the human resource development plan for TB laboratory?

☐ Yes

| If Yes, please specify when it was established | | |
| --- | --- | --- |
| ☐ Between 1994 and 2005 | ☐ Between 2006 and 2015 | ☐ Between 2016 and 2021 |
| ☐ Specify the exact year: | ☐ Specify the exact year: | ☐ Specify the exact year: |
| ☐ I do not know | | |

☐ No

☐ I donot know

**Q13**: when a new diagnostic technology is endorsed by WHO, for your country, how long does it take for TB laboratory personnel to be trained?

| ☐ 1 to 3 years | ☐ 3 to 5 years | ☐ ≥ 5 years |
| --- | --- | --- |

**Q14**: Is your National TB Reference Laboratory upgraded at level 3 of the biosafety (BSL-3 level or High-risk TB laboratory or containment laboratory) according to the WHO TB laboratory biosafety manual issued in 2012 available at [Tuberculosis laboratory biosafety manual (who.int)](https://www.who.int/publications/i/item/9789241504638)?

☐ Yes

| If Yes, please specify when it was established | | |
| --- | --- | --- |
| ☐ Between 1994 and 2005 | ☐ Between 2006 and 2015 | ☐ Between 2016 and 2021 |
| ☐ Specify the exact year: | ☐ Specify the exact year: | ☐ Specify the exact year: |
| ☐ I do not know | | |

☐ No

☐ I do not know

**Q15**: In 2007, WHO endorsed the use of two sputum specimens instead of three for screening of TB suspects. **W**hat is the **number of sputum specimens** to be examined for screening of TB cases in your country?

☐ Three

☐ Two

| If the response is “Two”, since when (year) did your country start the implementation of this policy? | | |
| --- | --- | --- |
| ☐ Between 2007 and 2009 | ☐ Between 2010 and 2011 | ☐ From 2012 and beyond |
| ☐ Specify the exact year: | ☐ Specify the exact year: | ☐ Specify the exact year: |
| ☐ I do not know | | |

If the response is “Two”, please select the appropriate response with regard to factors that contributed to the implementation

| **Enabling factors** | **Predisposing factors** | **Need factors** |
| --- | --- | --- |
| ☐ Availability of funding  ☐ Political commitment  ☐ In country partners  ☐ Winning grant  ☐ All of the above  ☐ None of the above  ☐ Others | ☐ Political security  ☐ Policy reform  ☐ Laboratory network assessment  ☐ Competent staff  ☐ Lab preparedness  ☐ Infrastructure upgrade  ☐ All of the above  ☐ None of the above  ☐ Others | ☐ Increase in TB Incidence  ☐ Increase in TB mortality  ☐ TB incidence and mortality  ☐ Emergency of DR-TB  ☐ All of the above  ☐ None of the above  ☐ Others |

**Q16**: in 2007, WHO recommended the use of automated **liquid culture** and **Drug Susceptibility Testing** (**DST**) as well as **rapid speciation**. How long did your country take to establish the following phases?

| **Liquid culture:** [Policy guidance on TB drug susceptibility testing (DST) of second-line drugs (SLD) (who.int)](https://www.who.int/news/item/23-07-2007-policy-guidance-on-tb-drug-susceptibility-testing-(dst)-of-second-line-drugs-(sld)) | **Rapid speciation** |
| --- | --- |
| a. Laboratory preparedness   ☐ 1 to 3 years  ☐ 3 to 5 years  ☐ ≥ 5 years   b. Technology transfer   ☐ 1 to 3 years  ☐ 3 to 5 years  ☐ ≥ 5 years   c. Routine testing and monitoring   ☐ 1 to 3 years  ☐ 3 to 5 years  ☐ ≥ 5 years  1) Do your guidelines recommend the use of 1st line DST (rifampicin and isoniazid) as the initial test for all RR-TB cases or for people at risk of DR-TB?  ☐Yes  ☐No | a. Laboratory preparedness   ☐ 1 to 3 years  ☐ 3 to 5 years  ☐ ≥ 5 years   b. Technology transfer   ☐ 1 to 3 years  ☐ 3 to 5 years  ☐ ≥ 5 years   c. Routine testing and monitoring   ☐ 1 to 3 years  ☐ 3 to 5 years  ☐ ≥ 5 years |

Please select the appropriate response with regard to factors that contributed to the implementation

| **Enabling factors** | **Predisposing factors** | **Need factors** |
| --- | --- | --- |
| ☐ Availability of funding  ☐ Political commitment  ☐ In country partners  ☐ Winning grant  ☐ All of the above  ☐ None of the above  ☐ Others | ☐ Political security  ☐ Policy reform  ☐ Laboratory network assessment  ☐ Competent staff  ☐ Lab preparedness  ☐ Infrastructure upgrade  ☐ All of the above  ☐ None of the above  ☐ Others | ☐ Increase in TB Incidence  ☐ Increase in TB mortality  ☐ TB incidence and mortality  ☐ Emergency of DR-TB  ☐ All of the above  ☐ None of the above  ☐ Others |

**Q17**: in 2008, WHO recommended the use of automated genotypic **Line Probe Assay for first line (1^st^ line LPA, updated in 2016) and 2^nd^ line phenotypic DST**. How long did your country take to establish the following phases?

| **Line Probe Assay for first line:** [policy_update_useofmolecular.indd (who.int)](https://apps.who.int/iris/bitstream/handle/10665/250586/9789241511261-eng.pdf;jsessionid=535FE67AD50F17A162A2B8CFCBFA0B75?sequence=1) | **2^nd^ line phenotypic DST:** [Policy guidance on drug-susceptibility testing (DST) of second-line antituberculosis drugs (who.int)](https://www.who.int/publications/i/item/WHO-HTM-TB-2008.392) |
| --- | --- |
| a. Laboratory preparedness   ☐ 1 to 3 years  ☐ 3 to 5 years  ☐ ≥ 5 years   b. Technology transfer   ☐ 1 to 3 years  ☐ 3 to 5 years  ☐ ≥ 5 years   c. Routine testing and monitoring   ☐ 1 to 3 years  ☐ 3 to 5 years  ☐ ≥ 5 years | a. Laboratory preparedness   ☐ 1 to 3 years  ☐ 3 to 5 years  ☐ ≥ 5 years   b. Technology transfer   ☐ 1 to 3 years  ☐ 3 to 5 years  ☐ ≥ 5 years   c. Routine testing and monitoring   ☐ 1 to 3 years  ☐ 3 to 5 years  ☐ ≥ 5 years |

Please select the appropriate response with regard to factors that contributed to the implementation

| **Enabling factors** | **Predisposing factors** | **Need factors** |
| --- | --- | --- |
| ☐ Availability of funding  ☐ Political commitment  ☐ In country partners  ☐ Winning grant  ☐ All of the above  ☐ None of the above  ☐ Others | ☐ Political security  ☐ Policy reform  ☐ Laboratory network assessment  ☐ Competent staff  ☐ Lab preparedness  ☐ Infrastructure upgrade  ☐ All of the above  ☐ None of the above  ☐ Others | ☐ Increase in TB Incidence  ☐ Increase in TB mortality  ☐ TB incidence and mortality  ☐ Emergency of DR-TB  ☐ All of the above  ☐ None of the above  ☐ Others |

**Q18**: in 2010, WHO recommended the use of the following automated diagnostic technologies below. How long did your country take to establish the following phases per each technology?

| **LED Microscopy:** [Fluorescent light-emitting diode (‎LED)‎ microscopy for diagnosis of tuberculosis: policy statement (who.int)](https://apps.who.int/iris/handle/10665/44602) | **Non commercial culture and DST :** [Non-commercial culture and drug-susceptibility testing methods for ... (yumpu.com)](https://www.yumpu.com/en/document/read/34277038/non-commercial-culture-and-drug-susceptibility-testing-methods-for-) | **Xpert MTB/RIF** in **adults** presumptive of MDR or **HIV associated TB:** [Automated real-time nucleic acid amplification technology for rapid and simultaneous detection of tuberculosis and rifampicin resistance: Xpert MTB/RIF system: policy statement (who.int)](https://apps.who.int/iris/handle/10665/44586) |
| --- | --- | --- |
| a. Laboratory preparedness   ☐ 1 to 3 years  ☐ 3 to 5 years  ☐ ≥ 5 years   b. Technology transfer   ☐ 1 to 3 years  ☐ 3 to 5 years  ☐ ≥ 5 years   c. Routine testing and monitoring   ☐ 1 to 3 years  ☐ 3 to 5 years  ☐ ≥ 5 years | a. Laboratory preparedness   ☐ 1 to 3 years  ☐ 3 to 5 years  ☐ ≥ 5 years   b. Technology transfer   ☐ 1 to 3 years  ☐ 3 to 5 years  ☐ ≥ 5 years   c. Routine testing and monitoring   ☐ 1 to 3 years  ☐ 3 to 5 years  ☐ ≥ 5 years | a. Laboratory preparedness   ☐ 1 to 3 years  ☐ 3 to 5 years  ☐ ≥ 5 years   b. Technology transfer   ☐ 1 to 3 years  ☐ 3 to 5 years  ☐ ≥ 5 years   c. Routine testing and monitoring   ☐ 1 to 3 years  ☐ 3 to 5 years  ☐ ≥ 5 years  Between 2010 and 2013 : Xpert MTB/RIF used as initial test for:  ☐ all adult suspected of TB  ☐ only risk groups (HIV/DR-TB)  ☐ all of the above  ☐ none of the above |

Please select the appropriate response with regard to factors that contributed to the implementation

| **Enabling factors** | **Predisposing factors** | **Need factors** |
| --- | --- | --- |
| ☐ Availability of funding  ☐ Political commitment  ☐ In country partners  ☐ Winning grant  ☐ All of the above  ☐ None of the above  ☐ Others | ☐ Political security  ☐ Policy reform  ☐ Laboratory network assessment  ☐ Competent staff  ☐ Lab preparedness  ☐ Infrastructure upgrade  ☐ All of the above  ☐ None of the above  ☐ Others | ☐ Increase in TB Incidence  ☐ Increase in TB mortality  ☐ TB incidence and mortality  ☐ Emergency of DR-TB  ☐ All of the above  ☐ None of the above  ☐ Others |

**Q19**: in 2013, WHO updated the policy and recommended the use of **Xpert MTB/RIF as initial diagnostic test for all people (adults and children) with signs and symptoms for TB**. How long did your country take to establish the following phases?

| a. Laboratory preparedness   ☐ 1 to 3 years  ☐ 3 to 5 years  ☐ ≥ 5 years | b. Technology transfer   ☐ 1 to 3 years  ☐ 3 to 5 years  ☐ ≥ 5 years | c. Routine testing and monitoring   ☐ 1 to 3 years  ☐ 3 to 5 years  ☐ ≥ 5 years  After 2013 :  1) Is Xpert MTB/RIF included in your policy as initial test for all TB suspected?  ☐ Yes  ☐ No  2) Is Xpert MTB/RIF used as initial TB Diagnostic test for:  ☐ adult and children suspected of TB  ☐ only adults suspected of TB  ☐ only risk groups (HIV/DR-TB)  ☐ all of the above |
| --- | --- | --- |

Please select the appropriate response with regard to factors that contributed to the implementation

| **Enabling factors** | **Predisposing factors** | **Need factors** |
| --- | --- | --- |
| ☐ Availability of funding  ☐ Political commitment  ☐ In country partners  ☐ Winning grant  ☐ All of the above  ☐ None of the above  ☐ Others | ☐ Political security  ☐ Policy reform  ☐ Laboratory network assessment  ☐ Competent staff  ☐ Lab preparedness  ☐ Infrastructure upgrade  ☐ All of the above  ☐ None of the above  ☐ Others | ☐ Increase in TB Incidence  ☐ Increase in TB mortality  ☐ TB incidence and mortality  ☐ Emergency of DR-TB  ☐ All of the above  ☐ None of the above  ☐ Others |

**Q20**:

a) In 2015, WHO updated the policy and recommended the use of **Lateral flow- Urine Lipoarabinomannan assay (LF-LAM):** [The use of lateral flow urine lipoarabinomannan assay (‎LF-LAM)‎ for the diagnosis and screening of active tuberculosis in people living with HIV: policy guidance (who.int)](https://apps.who.int/iris/handle/10665/193633?search-result=true&query=Lateral+flow-+Urine+Lipoarabinomannan+assay&scope=&rpp=10&sort_by=score&order=desc). How long did your country take to establish the following phases?

| a. Laboratory preparedness   ☐ 1 to 3 years  ☐ 3 to 5 years  ☐ ≥ 5 years | b. Technology transfer   ☐ 1 to 3 years  ☐ 3 to 5 years  ☐ ≥ 5 years | c. Routine testing and monitoring   ☐ 1 to 3 years  ☐ 3 to 5 years  ☐ ≥ 5 years |
| --- | --- | --- |

Please select the appropriate response with regard to factors that contributed to the implementation

| **Enabling factors** | **Predisposing factors** | **Need factors** |
| --- | --- | --- |
| ☐ Availability of funding  ☐ Political commitment  ☐ In country partners  ☐ Winning grant  ☐ All of the above  ☐ None of the above  ☐ Others | ☐ Political security  ☐ Policy reform  ☐ Laboratory network assessment  ☐ Competent staff  ☐ Lab preparedness  ☐ Infrastructure upgrade  ☐ All of the above  ☐ None of the above  ☐ Others | ☐ Increase in TB Incidence  ☐ Increase in TB mortality  ☐ TB incidence and mortality  ☐ Emergency of DR-TB  ☐ All of the above  ☐ None of the above  ☐ Others |

b) In 2019, WHO updated the policy and recommended the use of **Lateral flow- Urine Lipoarabinomannan assay (LF-LAM)** [Lateral flow urine lipoarabinomannan assay (‎LF-LAM)‎ for the diagnosis of active tuberculosis in people living with HIV: policy update 2019 (who.int)](https://apps.who.int/iris/handle/10665/329479?search-result=true&query=Lateral+flow-+Urine+Lipoarabinomannan+assay&scope=&rpp=10&sort_by=score&order=desc). How long did your country take to establish the following phases?

| a. Laboratory preparedness   ☐ 1 to 3 years  ☐ 3 to 5 years  ☐ ≥ 5 years | b. Technology transfer   ☐ 1 to 3 years  ☐ 3 to 5 years  ☐ ≥ 5 years | c. Routine testing and monitoring   ☐ 1 to 3 years  ☐ 3 to 5 years  ☐ ≥ 5 years |
| --- | --- | --- |

Please select the appropriate response with regard to factors that contributed to the implementation

| **Enabling factors** | **Predisposing factors** | **Need factors** |
| --- | --- | --- |
| ☐ Availability of funding  ☐ Political commitment  ☐ In country partners  ☐ Winning grant  ☐ All of the above  ☐ None of the above  ☐ Others | ☐ Political security  ☐ Policy reform  ☐ Laboratory network assessment  ☐ Competent staff  ☐ Lab preparedness  ☐ Infrastructure upgrade  ☐ All of the above  ☐ None of the above  ☐ Others | ☐ Increase in TB Incidence  ☐ Increase in TB mortality  ☐ TB incidence and mortality  ☐ Emergency of DR-TB  ☐ All of the above  ☐ None of the above  ☐ Others |

c) Is Urine LAM diagnostic algorithn in your country guidelines?

☐ Yes, if Yes, is it used in

☐ Adults

☐ Children and adolescents

☐ all hospitals

☐ limited number of hospitals

☐ all ART centers

☐ limited number of ART centers

☐ TB Programme

☐ HIV Programme

☐ No

**Q21**: in 2016, WHO updated the policy and recommended the use of **two diagnostic technologies below**. How long did your country take to establish the following phases per each technology?

| **2^nd^ line LPA:** [WHOPolicyStatementSLLPA.pdf](https://www.who.int/tb/WHOPolicyStatementSLLPA.pdf) | **TB LAMP:** [The use of loop-mediated isothermal amplification (‎TB-LAMP)‎ for the diagnosis of pulmonary tuberculosis: policy guidance (who.int)](https://apps.who.int/iris/handle/10665/249154) |
| --- | --- |
| a. Laboratory preparedness   ☐ 1 to 3 years  ☐ 3 to 5 years  ☐ ≥ 5 years   b. Technology transfer   ☐ 1 to 3 years  ☐ 3 to 5 years  ☐ ≥ 5 years   c. Routine testing and monitoring   ☐ 1 to 3 years  ☐ 3 to 5 years  ☐ ≥ 5 years  In your country:  1) Does the Policy recommends that all RIF-resistant by Xpert undergo 2nd line LPA?  ☐ Yes  ☐ No | a. Laboratory preparedness   ☐ 1 to 3 years  ☐ 3 to 5 years  ☐ ≥ 5 years   b. Technology transfer   ☐ 1 to 3 years  ☐ 3 to 5 years  ☐ ≥ 5 years   c. Routine testing and monitoring   ☐ 1 to 3 years  ☐ 3 to 5 years  ☐ ≥ 5 years  In your country:  1) Does the Policy recommend the implementation of TB-LAMP a test  to replace microscopy for diagnosing of TB?  ☐ Yes  ☐ No |

Please select the appropriate response with regard to factors that contributed to the implementation

| **Enabling factors** | **Predisposing factors** | **Need factors** |
| --- | --- | --- |
| ☐ Availability of funding  ☐ Political commitment  ☐ In country partners  ☐ Winning grant  ☐ All of the above  ☐ None of the above  ☐ Others | ☐ Political security  ☐ Policy reform  ☐ Laboratory network assessment  ☐ Competent staff  ☐ Lab preparedness  ☐ Infrastructure upgrade  ☐ All of the above  ☐ None of the above  ☐ Others | ☐ Increase in TB Incidence  ☐ Increase in TB mortality  ☐ TB incidence and mortality  ☐ Emergency of DR-TB  ☐ All of the above  ☐ None of the above  ☐ Others |

**Q22**: in 2017, WHO updated the policy and recommended the use of **Xpert Ultra**. How long did your country take to establish the following phases?

| a. Laboratory preparedness   ☐ 1 to 3 years  ☐ 3 to 5 years  ☐ ≥ 5 years | b. Technology transfer   ☐ 1 to 3 years  ☐ 3 to 5 years  ☐ ≥ 5 years | c. Routine testing and monitoring   ☐ 1 to 3 years  ☐ 3 to 5 years  ☐ ≥ 5 years  1) In your country, does the policy recommend transitioning from Xpert MTB/RIF to Ultra?  ☐ Yes  ☐ No |
| --- | --- | --- |

Please select the appropriate response with regard to factors that contributed to the implementation

| **Enabling factors** | **Predisposing factors** | **Need factors** |
| --- | --- | --- |
| ☐ Availability of funding  ☐ Political commitment  ☐ In country partners  ☐ Winning grant  ☐ All of the above  ☐ None of the above  ☐ Others | ☐ Political security  ☐ Policy reform  ☐ Laboratory network assessment  ☐ Competent staff  ☐ Lab preparedness  ☐ Infrastructure upgrade  ☐ All of the above  ☐ None of the above  ☐ Others | ☐ Increase in TB Incidence  ☐ Increase in TB mortality  ☐ TB incidence and mortality  ☐ Emergency of DR-TB  ☐ All of the above  ☐ None of the above  ☐ Others |

**Q23**: in your country, kindly specify the use and the location of the following technologies:

I. **Smear microscopy (ZN or LED)**

| **Use**  ☐ Primary diagnosis  ☐ Primary diagnosis of HIV negative cases  ☐ Primary diagnosis and Follow-Up  ☐ Follow-up only  **1)** In your country, does the policy recommend the use of LED instead of ZN microscopy ?  ☐ Yes  ☐ No | **Location**  ☐ Central level  ☐ Regional level  ☐ Peripherl level  ☐ All of the above |
| --- | --- |

Please select the appropriate response with regard to factors that contributed to the implementation

| **Enabling factors** | **Predisposing factors** | **Need factors** |
| --- | --- | --- |
| ☐ Availability of funding  ☐ Political commitment  ☐ In country partners  ☐ Winning grant  ☐ All of the above  ☐ None of the above  ☐ Others | ☐ Political security  ☐ Policy reform  ☐ Laboratory network assessment  ☐ Competent staff  ☐ Lab preparedness  ☐ Infrastructure upgrade  ☒ All of the above  ☐ None of the above  ☐ Others | ☐ Increase in TB Incidence  ☐ Increase in TB mortality  ☐ TB incidence and mortality  ☐ Emergency of DR-TB  ☐ All of the above  ☐ None of the above  ☐ Others |

II. **Xpert MTB/RIF or Ultra:** please select the appropriate response

| **Use**  ☐ as the initial diagnostic test for TB  ☐ as a follow-on test after smear microscopy  ☐ both | **Location**  ☐ Central level  ☐ Regional level  ☐ Peripherl level  ☐ All of the above |
| --- | --- |

Please select the appropriate response with regard to factors that contributed to the implementation

| **Enabling factors** | **Predisposing factors** | **Need factors** |
| --- | --- | --- |
| ☐ Availability of funding  ☐ Political commitment  ☐ In country partners  ☐ Winning grant  ☐ All of the above  ☐ None of the above  ☐ Others | ☐ Political security  ☐ Policy reform  ☐ Laboratory network assessment  ☐ Competent staff  ☐ Lab preparedness  ☐ Infrastructure upgrade  ☐ All of the above  ☐ None of the above  ☐ Others | ☐ Increase in TB Incidence  ☐ Increase in TB mortality  ☐ TB incidence and mortality  ☐ Emergency of DR-TB  ☐ All of the above  ☐ None of the above  ☐ Others |

III. **The 1st and 2^nd^ line LPA :** please select the appropriate response

| **The 1st line LPA** | | **The 2^nd^ line LPA** | |
| --- | --- | --- | --- |
| - **Level of the utilization**   ☐ Central level  ☐ Regional level  ☐ Both | - **Frequency of utilization**   ☐ Weekly  ☐ Bi-weekly  ☐ Monthly | - **Level of the utilization**   ☐ Central level  ☐ Regional level  ☐ Both  1) Do your guidelines recommend the use of 2nd line LPA as the initial test for people with confirmed RR- and MDR-TB?  ☐Yes  ☐No | - **Frequency of utilization**   ☐ Weekly  ☐ Bi-weekly  ☐ Monthly |

Please select the appropriate response with regard to factors that contributed to the implementation

| **Enabling factors** | **Predisposing factors** | **Need factors** |
| --- | --- | --- |
| ☐ Availability of funding  ☐ Political commitment  ☐ In country partners  ☐ Winning grant  ☐ All of the above  ☐ None of the above  ☐ Others | ☐ Political security  ☐ Policy reform  ☐ Laboratory network assessment  ☐ Competent staff  ☐ Lab preparedness  ☐ Infrastructure upgrade  ☐ All of the above  ☐ None of the above  ☐ Others | ☐ Increase in TB Incidence  ☐ Increase in TB mortality  ☐ TB incidence and mortality  ☐ Emergency of DR-TB  ☐ All of the above  ☐ None of the above  ☐ Others |

IV. **The 1^st^ and 2^nd^ line phenotypic DST**

| **The 1^st^ line phenotypic DST** | | **The 2^nd^ line phenotypic DST** | |
| --- | --- | --- | --- |
| - **Level of the utilization**   ☐ Central level  ☐ Regional level  ☐ Both  1) Do your guidelines recommend that all bacteriologically confirmed TB cases undergo lst line DST?  ☐ Yes  ☐ No | - **Frequency of utilization**   ☐ Weekly  ☐ Bi-weekly  ☐ Monthly | - **Level of the utilization**   ☐ Central level  ☐ Regional level  ☐ Both  1) Do your guidelines recommend that all RR par Xpert and MDR-TB cases undergo 2nd line DST?  ☐ Yes  ☐ No | - **Frequency of utilization**   ☐ Weekly  ☐ Bi-weekly  ☐ Monthly |

Please select the appropriate response with regard to factors that contributed to the implementation

| **Enabling factors** | **Predisposing factors** | **Need factors** |
| --- | --- | --- |
| ☐ Availability of funding  ☐ Political commitment  ☐ In country partners  ☐ Winning grant  ☐ All of the above  ☐ None of the above  ☐ Others | ☐ Political security  ☐ Policy reform  ☐ Laboratory network assessment  ☐ Competent staff  ☐ Lab preparedness  ☐ Infrastructure upgrade  ☐ All of the above  ☐ None of the above  ☐ Others | ☐ Increase in TB Incidence  ☐ Increase in TB mortality  ☐ TB incidence and mortality  ☐ Emergency of DR-TB  ☐ All of the above  ☐ None of the above  ☐ Others |

**Q24**: please select the main impediments and barriers for a rapid implementation of TB diagnostic technologies in your country

☐ inadequate human resource

☐ procurement issues

☐ poor electricity- and water- supply

☐ insufficient finance to procure the test

☐ Lack of awareness of the technology

☐ Biosafety issues

☐ The test is only relevant for a small population

☐ The test is not in the TB programme's mandate

☐ Health-care workers are not enough trained to identify and evaluate eligible patients

☐ I am unaware of the test existence

☐ The test is not approved/registered by the national regulatory agency

☐ Pilot studies on the test in the country are still to be evaluated

☐ Unsure of how many tests are needed in the country

☐ Concerns about additional staff workload

☐ Concerns about non-compliance with other TB diagnostic test

☐ Lack of agreement by policy-makers on using the test

☐ Clinicians do not value the test and will treat TB empirically

End
